# MASLD prevalence by ultrasonography and clinical profile in obese adults attending a Mexican primary care unit: a cross-sectional study

**DOI:** 10.64898/2026.07.13.26357943

**Authors:** Efren Rene Nieves-Ruiz, Victor Junior Godinez-Vazquez, Ricardo Arguello-Flores, Alejandro Aguilar-Cruz, Florencia Monserrat Belman-Estrada

## Abstract

**Background:** Metabolic dysfunction-associated steatotic liver disease (MASLD) is linked to obesity and cardiometabolic dysfunction, yet local prevalence data from Mexican first-level care units are scarce. We estimated the prevalence of MASLD by B-mode ultrasonography and characterized the clinical profile of obese adults in a family medicine unit in Leon, Mexico.

**Methods:** Cross-sectional study of 55 adults with obesity (body mass index [BMI] ≥30 kg/m^2^) registered at one medical office of a Mexican Social Security Institute family medicine unit (April–October 2024). Hepatic steatosis was graded by B-mode ultrasonography; MASLD was defined as ultrasonographic steatosis plus at least one cardiometabolic criterion (all participants met the obesity criterion). Prevalence was reported with Wilson 95% confidence intervals (CIs); associations with obesity grade were assessed by Cochran-Armitage trend and Fisher exact tests.

**Results:** MASLD was present in 37 of 55 participants (67.3%; 95% CI, 54.1–78.2). Mild (grade 1) steatosis predominated (45.5%). Prevalence increased across obesity grades—59.0%, 75.0%, and 100% for grades 1, 2, and 3, respectively (trend P=0.022). Type 2 diabetes and hypertension each affected 50.9% of participants; the sample was predominantly female (74.5%).

**Conclusion:** Two of every three obese adults screened in primary care had ultrasonographic MASLD, with a significant rise across obesity grades. Opportunistic ultrasonography in obese primary-care patients may enable earlier MASLD detection.

## Introduction

NAFLD (non-alcoholic fatty liver disease) has been renamed after many medical societies debated changing the name and reached an international consensus on the term MASLD (metabolic dysfunction-associated steatotic liver disease). This change reflects the evolution of understanding of the disease and its metabolic implications, raising awareness of its clinical importance while minimizing the stigma associated with the terms “fatty” and “non-alcoholic,” and preserving continuity with the existing body of research on natural history, biomarkers, and clinical trials. Patients previously diagnosed with NAFLD are fully captured by the MASLD criteria, which can accelerate the development of new biomarkers and therapeutic drug development [1].

MASLD has been recognized as a more effective indicator for identifying metabolic dysfunction that increases risk in patients [2]. Eliminating the alcohol stigma allows recognition of the role of obesity, sedentary lifestyle, and energy-dense diets that promote adiposity and lipotoxicity.

MASLD not only represents liver damage and metabolic alterations; current evidence suggests associations with extrahepatic manifestations such as cognitive impairment that diminishes mental health and cognitive performance [3], as well as an increased risk of muscle mass loss associated with metabolic dysfunction, as demonstrated in a Korean population with asymptomatic MAFLD. This clarifies the role of metabolic dysfunction in muscle tissue function. Although low muscle strength was correlated with a higher risk of hepatic steatosis, the mechanisms underlying the adiposity–muscle interaction remain under investigation, with complex interactions arising [4,5].

The global prevalence estimated among overweight and obese adults was approximately 50.7%. Males reported a higher prevalence, estimated at 59%, while female prevalence was approximately 47.5%. Comorbidities such as type 2 diabetes appeared in 19.7% of cases and metabolic syndrome in 57.7%. Ultrasound was the most commonly used diagnostic technique, accounting for 51.3% of reported diagnoses [6].

In patients with a previous diagnosis of type 2 diabetes (T2D), a meta-analysis showed a global prevalence of 65.3%, with the highest prevalence in Eastern Europe (80.62%) and the Middle East (71.24%), and the lowest in Africa (53.1%). Fibrosis was present in 66.44% of cases confirmed by biopsy. These data demonstrate that T2D contributes to a progressive accumulation of metabolic complications over time, and that the coexistence of MAFLD and T2D represents a higher risk for the development of advanced fibrosis [7,8].

In the United States, steatohepatitis is estimated to be the fastest-growing cause of hepatocellular carcinoma and the principal indication for liver transplantation [9], representing a substantial economic burden and a public health concern.

Mexico reports one of the highest prevalences in the world, at 41.3% in the adult population, of which 40% have fibrosis. The predominant characteristics were a sedentary lifestyle and poor dietary habits. Male sex, obesity, metabolic syndrome, and elevated ALT were identified as predictors. In Mexican adolescents, the prevalence reported by ultrasound diagnosis was 69.6%, which was associated with a diagnosis of prediabetes [10,11].

The pathophysiology of MASLD shows that when the mechanisms regulating hepatic insulin sensitivity are affected by liver adiposity, hormones such as adiponectin and leptin, which are essential for regulating insulin response and influencing peripheral tissues, become dysregulated. Hepatic apoptosis induced by caspases plays an important role in disease progression. Hepatocytes generate signals that produce immunological reactions with cellular infiltration, creating a proinflammatory intrahepatic and extrahepatic condition. The multiple-hit theory is attributed to its pathogenesis, with factors such as microbiome dysbiosis interacting with epigenetic changes and alterations in hepatic lipid metabolism. Excessive dietary fat intake plays a significant role, creating conditions for lipotoxicity. When leptin resistance appears, GLP-1 can restore this sensitivity and promote satiety. A more comprehensive understanding of the mechanisms underlying lipotoxicity, oxidative stress, insulin resistance, and inflammation is necessary to advance the understanding of its pathophysiology and the development of treatment strategies [12–14].

## Diagnostic criteria

Diagnosis of MASLD requires documentation of hepatic steatosis (by imaging, validated biomarkers, or liver biopsy) plus the presence of at least one of the following cardiometabolic risk factors [1]:

- Overweight or obesity (BMI ≥25 kg/m^2^; ≥23 kg/m^2^ in Asian populations), or increased waist circumference (>94 cm in men / >80 cm in women)
- Fasting serum glucose ≥100 mg/dL (5.6 mmol/L), 2-hour post-load glucose ≥140 mg/dL (7.8 mmol/L), HbA1c ≥5.7%, type 2 diabetes, or ongoing antidiabetic treatment
- Blood pressure ≥130/85 mmHg, or ongoing antihypertensive treatment
- Plasma triglycerides ≥150 mg/dL (1.70 mmol/L), or lipid-lowering treatment
- Plasma HDL-cholesterol ≤40 mg/dL (1.0 mmol/L) in men / ≤50 mg/dL (1.3 mmol/L) in women, or lipid-lowering treatment

Resmetirom is the first drug specifically approved by the FDA to target hepatic steatosis, acting as an enhancer of hepatic metabolism [15]. Resmetirom (Rezdiffra) was developed under the accelerated drug approval pathway to address unmet medical needs in patients with this serious condition. Approval was based on the results of a randomized clinical trial that demonstrated significant improvement in liver scarring assessed by biopsy. Its mechanism of action involves agonism of the thyroid hormone receptor-beta (THR-β), which modulates hepatic lipid metabolism by reducing liver enzymes and fatty acids, with improvement in hepatic fibrosis and without affecting systemic thyroid function. The most common adverse effects are diarrhea and nausea. Interactions with other medications, such as statins, should be monitored [16,17].

Preliminary randomized clinical trial results suggest that acetylsalicylic acid at a daily dose of 81 mg for 6 months reduces hepatic fat content; however, further studies are needed to confirm these findings, which open a new therapeutic avenue [18].

Future research on pharmacological treatment will focus on combination therapy employing drugs with different targets, reflecting the complex pathophysiology of the metabolic condition, with the goal of maintaining sustained benefits over time under close safety monitoring [19].

At present, lifestyle modification remains the first-line approach to treating MASLD, although pharmacological agents that support weight loss can improve the symptoms associated with adiposity and lipotoxicity [20]. Lipidomic analysis of cholesterol-induced DNA damage suggests that senescent macrophages act as key drivers of inflammation and that their accumulation contributes to fibrosis. Targeting senescent macrophages may therefore represent a future therapeutic strategy to reduce inflammation in MASLD and yield novel biomarkers and therapeutic targets aimed at reducing steatosis and the hepatic lipid dysregulation driven by aging hepatic cells [21].

The aims of this study were to estimate the prevalence of MASLD by ultrasound in patients with risk factors and to characterize the clinical and sociodemographic profile of affected patients in a family medicine setting at a primary care unit in León, Guanajuato, Mexico.

## Methods

A cross-sectional, observational study included 55 adults with obesity registered at medical office No. 2 (morning shift) of Family Medicine Unit No. 56 of the Mexican Social Security Institute (IMSS) in León, Guanajuato, Mexico. The source population comprised all adults with obesity with active registration assigned to this medical office, totaling 1,400. The study was designed and reported in accordance with the STROBE (Strengthening the Reporting of Observational Studies in Epidemiology) guideline for cross-sectional studies.

The sample size was calculated using the formula for estimation of a single proportion, with a 95% confidence level, a precision of 5%, and an expected proportion of 5%, yielding a required sample of 69 patients (with finite-population correction).

General data (age and sex), comorbidities, and anthropometric measurements (weight, height, and BMI) were recorded. Weight and height were measured with a calibrated scale and stadiometer, and BMI was used to classify obesity grade.

Inclusion criteria were adults aged 35–65 years with current registration at the study medical office and BMI ≥30 kg/m^2^, or adults presenting with initial symptoms suggestive of MASLD (asthenia, adynamia, acanthosis nigricans, or long-standing right upper-quadrant pain after exclusion of alternative gallbladder pathology). Exclusion criteria were a previous diagnosis of any degree of hepatic steatosis, current nutritional follow-up, age younger than 35 years, and absence of current registration at the study medical office.

Hepatic steatosis was assessed by the Department of Radiology and Imaging using B-mode ultrasonography and a stratified steatosis grading scale (grade 1, mild; grade 2, moderate; grade 3, severe). B-mode ultrasonography detects hepatic steatosis rather than MASLD as such; in accordance with the 2023 multisociety nomenclature, ultrasonographic steatosis was classified as MASLD when accompanied by at least one cardiometabolic risk factor.

Because every participant met the obesity criterion (BMI ≥30 kg/m^2^), all participants with ultrasonographic steatosis satisfied the MASLD definition.

The study period was April to October 2024. A non-probability convenience sampling method was used; of the 69 patients initially recruited, 55 completed the study.

### Statistical analysis

Continuous variables were summarized as mean and range, and categorical variables as absolute frequencies and percentages. The prevalence of MASLD was reported with 95% CIs calculated by the Wilson method. The association between MASLD and obesity grade (an ordinal exposure) was assessed with the Cochran-Armitage test for trend; the dichotomized comparison (obesity grade ≥2 vs. grade 1) was additionally examined with the Fisher exact test, and prevalence and odds ratios were reported with 95% CIs. Descriptive statistics were computed in IBM SPSS Statistics ver. 22.0 (IBM Corp., Armonk, NY, USA); trend and exact tests and interval estimates were computed in Python ver. 3 (SciPy). A two-sided P<0.05 was considered statistically significant.

### Ethics statement

This study was conducted in accordance with the principles of the Declaration of Helsinki. The protocol was reviewed and approved by the Local Health Research and Ethics Committee (Comité Local de Investigación en Salud No. 1005) of the Mexican Social Security Institute (IMSS) on March 4, 2024 (institutional registration No. R-2024-1005-008; COFEPRIS registry 17 CI 11 020 031; CONBIOÉTICA registry 11 CEI 004 20190709). The study was classified as minimal-risk research under Article 17 of the Regulations of the Mexican General Health Law on Health Research. Written informed consent was obtained from all participants.

### Use of AI-assisted technologies

The authors used a generative artificial intelligence tool (Anthropic Claude) to assist with statistical computation, figure preparation, and language editing; all outputs were reviewed and verified by the authors, who take full responsibility for the content.

## Results

A total of 55 adults with obesity were evaluated. Females comprised the majority of the sample (74.5%, n=41) versus males (25.5%, n=14). The mean age was 49 years (range, 35– 64 years), and the most frequent age group was 56–60 years (29.1%, n=16); three participants (5.5%) were aged 60–65 years, all without MASLD.

Anthropometric analysis showed a mean BMI of 34.64 kg/m^2^ (range, 30.0–61.51 kg/m^2^). Obesity was grade 1 in 39 participants (70.9%), grade 2 in 8 (14.5%), and grade 3 in 8 (14.5%). Type 2 diabetes mellitus was present in 28 participants (50.9%; 24 controlled, 4 uncontrolled) and arterial hypertension in 28 (50.9%; 23 controlled, 5 uncontrolled); dyslipidemia was present in 14 participants (25.5%) (Table 1).

**Table 1.**
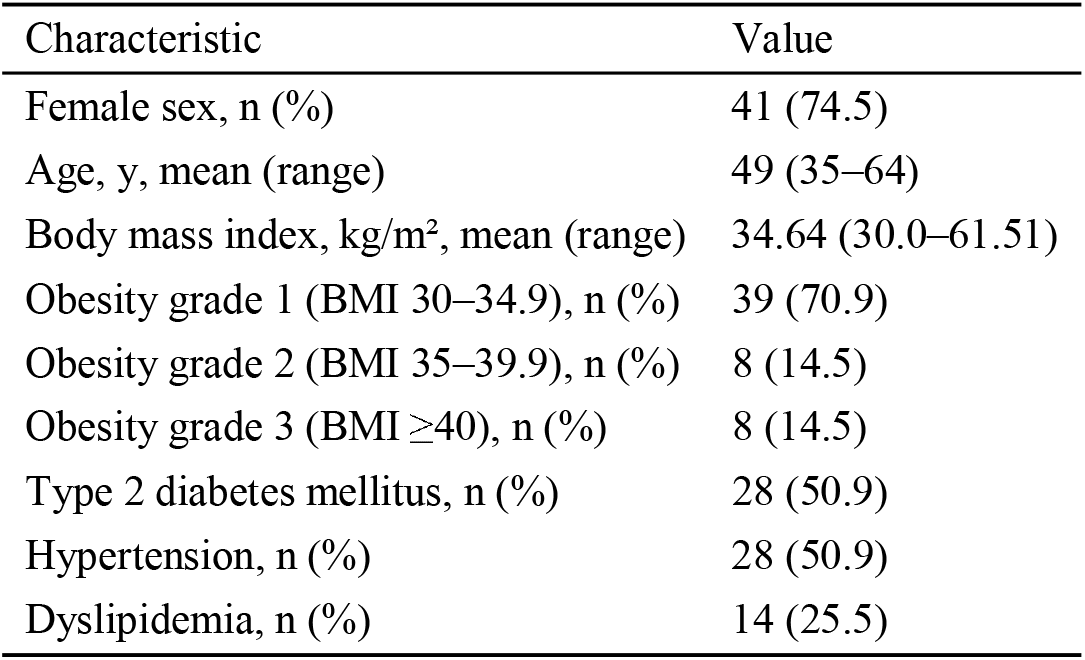
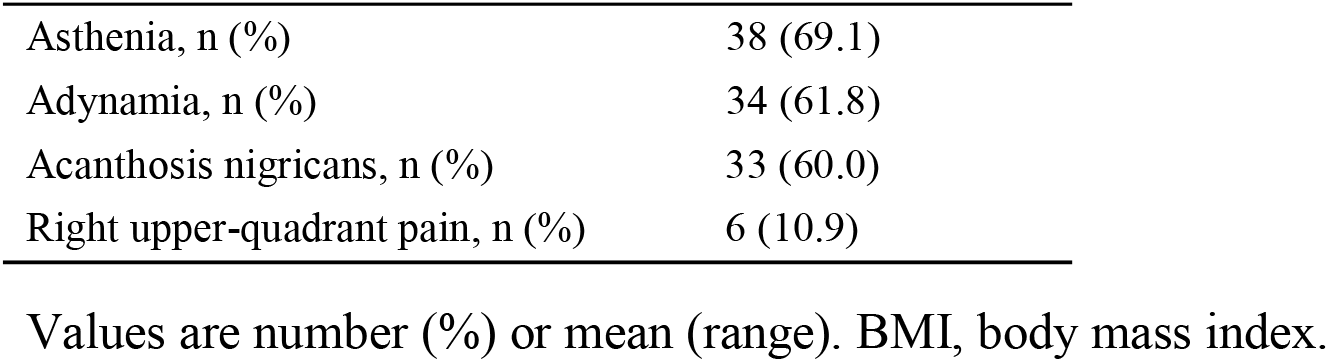
Baseline characteristics of the 55 obese participants.

Regarding clinical features, asthenia was the most frequently reported symptom (69.1%, n=38), followed by adynamia (61.8%, n=34) and acanthosis nigricans (60.0%, n=33); right upper-quadrant pain was the least frequent (10.9%, n=6). No participant presented with jaundice or hepatomegaly.

B-mode ultrasonography revealed abnormalities in 67.3% (n=37) of cases, whereas 32.7% (n=18) showed no abnormality. Among those with abnormalities, increased hepatic echogenicity was present in all cases; hepatic vascular structures were visible in 27 patients (49.1%) and the diaphragm in 36 (65.5%). The most frequent fat-distribution pattern was diffuse (65.5%, n=36), followed by no discernible pattern (32.7%, n=18) and multifocal (1.8%, n=1).

MASLD was diagnosed in 37 of 55 participants (67.3%; 95% CI, 54.1–78.2). Steatosis was graded as grade 1 (mild) in 25 participants (45.5%), grade 2 (moderate) in 11 (20.0%), and grade 3 (severe) in 1 (1.8%); 18 participants (32.7%) had no MASLD (Figure 3).

MASLD prevalence increased progressively across obesity grades: 59.0% (23/39; 95% CI, 43.4–72.9) in grade 1, 75.0% (6/8; 95% CI, 40.9–92.9) in grade 2, and 100% (8/8; 95% CI, 67.6–100) in grade 3—a statistically significant trend (Cochran-Armitage P=0.022) (Table 2, Figure 2). In the dichotomized comparison, participants with grade ≥2 obesity had a higher MASLD prevalence than those with grade 1 (87.5% vs. 59.0%; prevalence ratio, 1.48; 95% CI, 1.08–2.04; Fisher exact P=0.058). Among MASLD cases, grade 1 obesity accounted for the largest subgroup (62.1%, n=23). MASLD prevalence did not differ by sex (females, 68.3% [28/41]; males, 64.3% [9/14]; Fisher exact P=1.00).

**Table 2.**
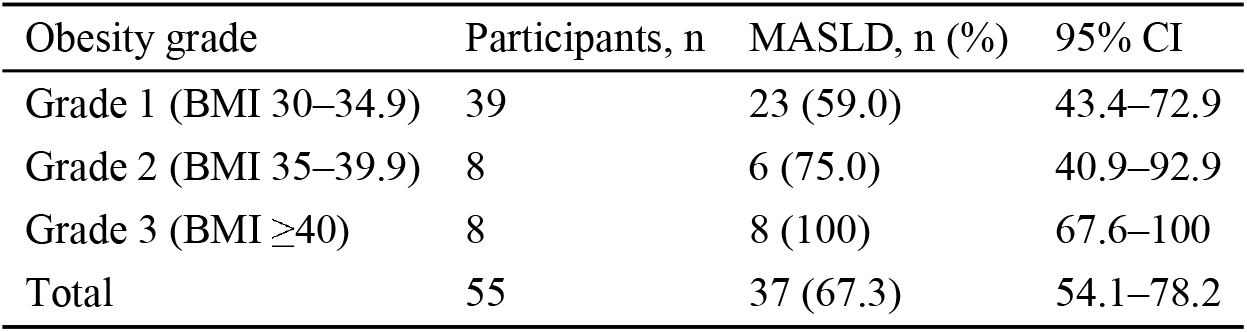
MASLD prevalence by obesity grade.

**Figure 1.**
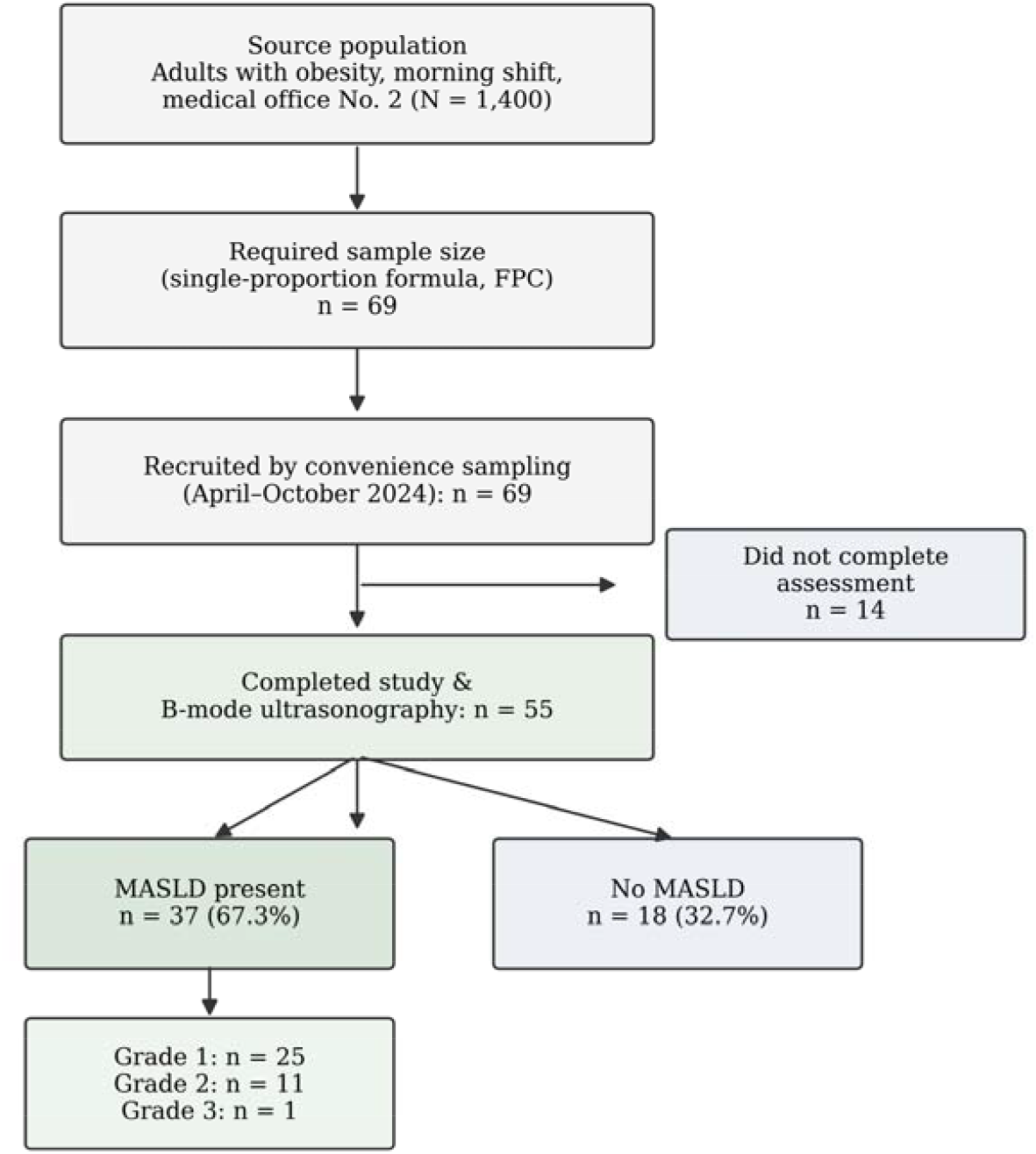
Participant flow (STROBE). Cascade from the source population through sample-size calculation, convenience recruitment, and study completion to MASLD ascertainment by B-mode ultrasonography. FPC, finite-population correction.

**Figure 2.**
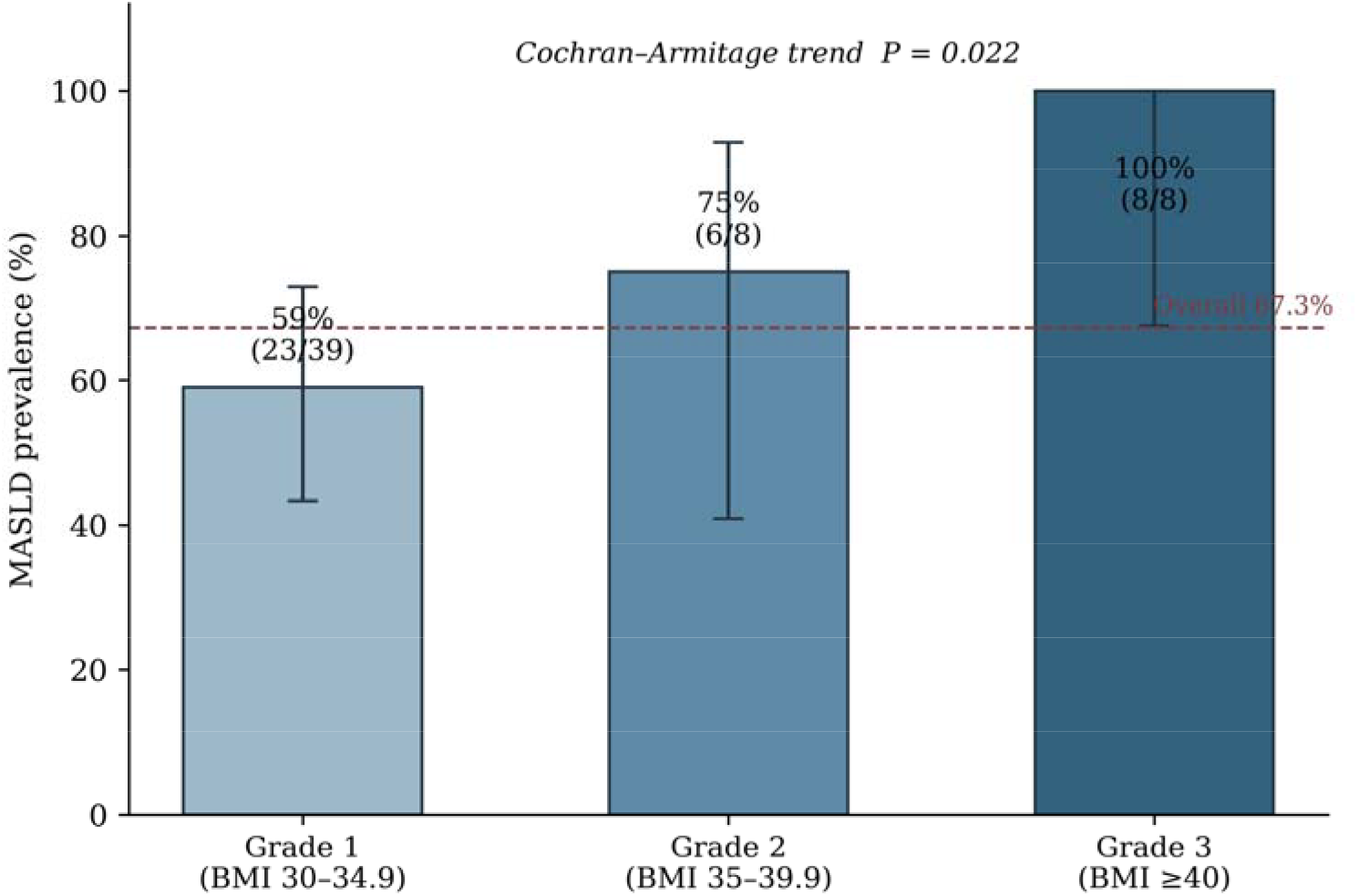
MASLD prevalence by obesity grade, shown with Wilson 95% confidence intervals. Prevalence increased across grades 1 to 3 (Cochran-Armitage test for trend, P=0.022); the dashed line marks the overall prevalence (67.3%).

**Figure 3.**
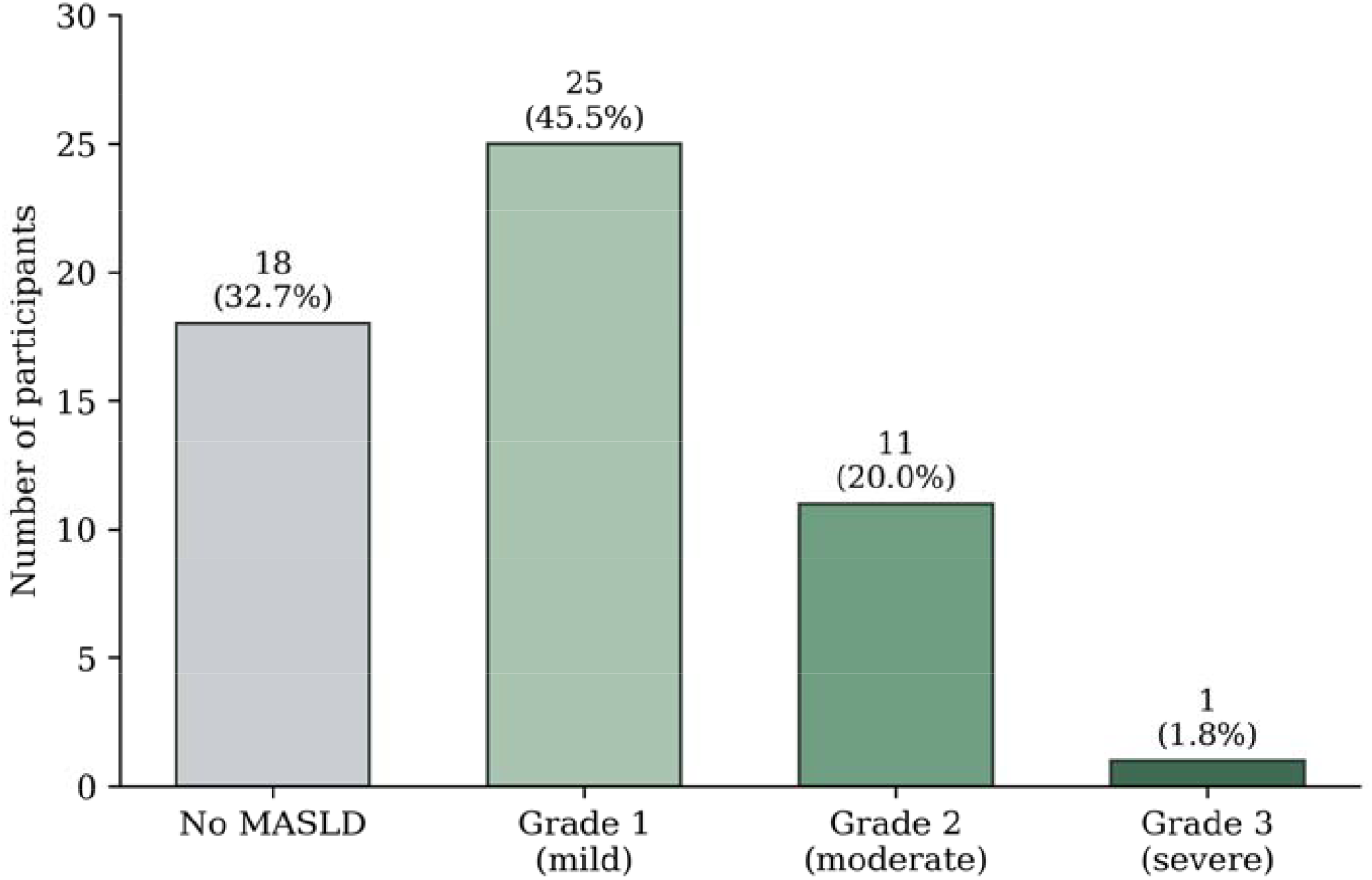
Distribution of MASLD status and steatosis severity among the 55 obese participants.

Prevalence estimates are shown with Wilson 95% confidence intervals. Cochran-Armitage test for trend across grades, P=0.022; dichotomized comparison (grade ≥2 vs. grade 1) by the Fisher exact test, prevalence ratio 1.48 (95% CI, 1.08–2.04), P=0.058. CI, confidence interval; MASLD, metabolic dysfunction-associated steatotic liver disease.

Additional ultrasonographic findings included chronic cholelithiasis in 9.1% (n=5), simple hepatic cysts in 7.3% (n=4), and—in 1.8% (n=1) each—calcifications, granular parenchymal changes, post-cholecystectomy status, and biliary sludge.

## Discussion

The most important finding of this study was the identification of metabolic dysfunction-associated steatotic liver disease (MASLD) by B-mode ultrasonography in 67.3% (n=37) of participants. Among these, grade 1 (mild) steatosis predominated at 45.5% (n=25), followed by grade 2 in 20.0% (n=11) and grade 3 in 1.8% (n=1). This distribution is comparable to that reported by Mohan et al. in a 2025 cross-sectional study of 110 participants undergoing concurrent computed tomography and ultrasonography assessment, in which grade I (mild) steatosis was likewise the most frequent ultrasonographic finding (36.4%), followed by grade II (31.8%) and grade III (13.6%). The authors also demonstrated a relationship between body mass index (BMI) and steatosis severity, with the obese subgroup showing the highest prevalence of severe (grade III) steatosis (46.7%), whereas no cases of severe disease were observed among underweight participants [22].

The relationship between adiposity and steatosis severity is consistent with the predominance of mild-to-moderate disease observed in our sample, in which grade 1 obesity (BMI 30–34.9 kg/m^2^) accounted for the largest subgroup of MASLD cases (62.1%, n=23). Our study population is restricted to patients with established obesity (mean BMI 34.64 kg/m^2^), distinguishing it from the population studied by Mohan et al. (mean BMI 27.8 kg/m^2^, predominantly overweight). The convergent finding that mild steatosis predominates across both studies supports the clinical utility of ultrasonography for early MASLD detection, particularly in metabolically at-risk populations where early diagnosis can optimize prognosis.

In our study of 55 obese patients, females comprised 74.5% (n=41) of the sample, with a mean age of 49 years and the highest representation in the 56–60 year age group (29.1%, n=16). This female predominance is consistent with the findings of Elnasieh et al. in a cross-sectional study of 292 patients with type 2 diabetes mellitus at King Saud Medical City in Riyadh, Saudi Arabia, in which MASLD prevalence was higher among females (61.0%) than among males (47.1%, *P* = 0.017) [23]. Our study did not perform sex-stratified analysis of MASLD prevalence due to the small number of male participants (n=14); nevertheless, the marked female predominance in our sample reflects a population profile in which female sex itself may contribute to MASLD risk. Jancova et al., in a 2025 review on MASLD and women’s health, reported that female sex hormones modulate MASLD risk and that menopause is associated with a more than twofold increase in MASLD prevalence, attributable to the loss of the protective metabolic effects of estrogen and the redistribution of adipose tissue toward visceral depots [24]. The age distribution observed in our sample, with a peak in the perimenopausal-to-postmenopausal range, is consistent with this hormonal transition as a contributing factor to the female predominance documented.

The mean BMI in our sample was 34.64 kg/m^2^ (range 30.0–61.51 kg/m^2^), corresponding to a population entirely within the obesity range, with grade 1 obesity (BMI 30–34.9 kg/m^2^) representing the largest subgroup. MASLD was diagnosed in 67.3% of participants, of whom grade 1 obesity accounted for 62.1% (n=23) of cases. These findings correspond to the relationship between BMI and MASLD prevalence reported by Elnasieh et al., who documented MASLD prevalence rates of 64.6% in patients with BMI 30–34.9 kg/m^2^, 75.8% in those with BMI 35–39.9 kg/m^2^, and 80.0% in patients with BMI ≥40 kg/m^2^ (*P* < 0.001) [23]. The progressive increase in MASLD prevalence with each obesity class supports the central role of adiposity as a driver of hepatic steatosis. Latif and Ahsan, in a cross-sectional study specifically evaluating obese patients with type 2 diabetes mellitus, similarly reported a high MASLD prevalence in this comorbid phenotype, reinforcing that the combination of obesity and dysglycemia substantially elevates the risk of steatotic liver disease [25]. The coexistence of cardiometabolic risk factors such as type 2 diabetes, hypertension, and dyslipidemia in our sample is consistent with the established conceptualization of MASLD as the hepatic manifestation of metabolic syndrome, in which obesity and insulin resistance act as interrelated drivers of disease [23,25].

Considering the American Heart Association presidential advisory on Cardiovascular-Kidney-Metabolic health as a key impact factor for cardiovascular mortality [26], MASLD should be considered a priority for screening and clinical recognition in patients with obesity and cardiovascular risk factors. Thus the presence of obesity, together with metabolic and cardiovascular risk factors, represents an opportunity for the early detection of MASLD in primary care.

MASLD prevalence in our sample rose monotonically across obesity grades (59.0%, 75.0%, and 100% for grades 1, 2, and 3; trend P=0.022), providing a quantitative, internally graded confirmation of the adiposity–steatosis relationship discussed above and mirroring the class-dependent gradient reported by Elnasieh et al. [23].

This study has several limitations. Its cross-sectional design precludes inferences about incidence or causality. The non-probability convenience sample drawn from a single medical office limits generalizability, and the modest sample size (n=55) yields wide confidence intervals, particularly within obesity-grade strata (notably grade 3, where all eight participants had MASLD). MASLD was defined by B-mode ultrasonography, which is operator-dependent and less sensitive than magnetic resonance–based methods for mild steatosis, and no histological confirmation was obtained. Finally, the marked female predominance may limit applicability to male populations.

In conclusion, two of every three obese adults screened in this primary care setting had ultrasonographic MASLD, with prevalence increasing significantly across obesity grades. These findings support opportunistic ultrasonographic screening for MASLD in obese primary-care patients with cardiometabolic risk factors, where early detection can inform timely lifestyle and metabolic intervention.

## Data Availability

All data produced in the present study are available upon reasonable request to the authors.

